# Physiological and perceptual effects of GLP-1 receptor agonists during alcohol consumption in people with obesity: a pilot study

**DOI:** 10.1101/2025.04.25.25326413

**Authors:** Fatima Quddos, Mary Fowler, Ana Carolina de Lima Bovo, Zacarya Elbash, Allison N. Tegge, Kirstin M. Gatchalian, Anita S. Kablinger, Alexandra G. DiFeliceantonio, Warren K. Bickel

## Abstract

Any increase in alcohol use is associated with an increase in risk of illness and mortality and consequences of chronic alcohol use include cancer, hypertension, heart and liver disease, and Alcohol Use Disorder. Glucagon-like peptide-1 receptor agonists (GLP-1RAs) are effective anti-glycemic and weight-loss medications with a strong safety record. There is substantial preclinical evidence and mounting retrospective and prospective randomized controlled trial evidence that GLP-1RAs could be effective for reducing alcohol consumption. However, the mechanism by which GLP-1RAs reduce alcohol intake remains unclear. While medications that reduce alcohol intake such as naltrexone and acamprosate have central nervous system action, disulfiram reduces alcohol intake through peripheral mechanisms. Here, we test whether GLP-1RAs alter alcohol’s peripheral pharmacokinetics as a potential mechanism of action for their alcohol intake suppressive effects. In this pilot study, twenty participants with obesity in the GLP-1RA or control group consumed a challenge dose of alcohol, and we measured breath alcohol (BrAC) and the subjective effects of alcohol. We observed a delayed rise in BrAC and subjective effects in the GLP-1RA group as compared to controls, that was not explained by nausea. These data provide preliminary evidence that GLP-1RAs could act through peripheral mechanisms to suppress alcohol intake.

## Introduction

More than half of US adults currently use alcohol^1^. Alcohol use has negative immediate and long-term health consequences. A single episode of excessive drinking can raise the risk of injuries, violence, and alcohol poisoning, among other harmful outcomes^2^. Alcohol is considered a Group 1 carcinogen, causally linked to seven types of cancer^3^. Indeed, in January 2025, the Surgeon General of the United States released a report raising awareness of the links between cancer and alcohol consumption^4^. Any increase in alcohol use is associated with an increase in risk of illness and mortality. Long-term consequences of chronic alcohol use also include other health-related illnesses, such as high blood pressure, heart and liver disease, and Alcohol Use Disorder^2^. In the US, 178,000 deaths per year are attributable to alcohol use and increases are projected. Both alcohol use and alcohol related deaths increased in the US during the COVID-19 pandemic^5,6^. Given these trends and the risks associated with any alcohol use, treatments that decrease alcohol use even in people not meeting criteria for alcohol use disorder, have the potential to improve health.

Glucagon-like peptide-1 receptor agonists (GLP-1RAs) are effective anti-glycemic and weight-loss medications with a strong safety record. Preclinical studies with various types of GLP-1RAs have demonstrated reductions in alcohol intake^7–15^. In humans, retrospective cohort studies have found people taking GLP-1RAs are less likely to develop alcohol use disorder (AUD)^16^ and less likely to be hospitalized for acute alcohol intoxication^17^. Participants taking GLP-1RAs semaglutide or tirzepatide in a retrospective survey study, reported reduced drinking and binge drinking episodes^18^. GLP-1RA exenatide administered once a week decreased heavy drinking days and total alcohol consumption, but only for a subset of participants with body mass index (BMI) over 30^19,20^. A secondary analysis of a randomized controlled trial (RCT) for smoking cessation found GLP-1RA dulaglutide decreased alcohol intake by 36%^21^. Another GLP-1RA, semaglutide, once weekly, reduced alcohol consumption in a challenge task and drinks per drinking day^22^. Overall, there is substantial preclinical evidence and mounting retrospective and prospective RCT evidence that GLP-1RAs could be effective for reducing alcohol consumption.

The mechanism by which GLP-1RAs reduce alcohol intake is not well understood.

However, like all drugs with addictive potential, alcohol must reach the brain to exert its effects. Alcohol enters the blood in the upper intestine and crosses the blood brain barrier where it acts in the central nervous system as a gamma-aminobutyric acid A (GABA-A) receptor agonist, among other targets^23,24^. Therefore, possible mechanisms for the effects of GLP-1RA involve central nervous system effects or peripheral effects on gastric emptying, as alcohol must reach the upper intestine on its path to the brain. Survey data demonstrate people taking GLP-1RAs semaglutide and tirzepatide report reduced sedative and stimulative effects of alcohol^18^. A scrape and analysis of the social media platform reddit revealed reduced craving and increased negative effects of drinking alcohol ^18^. Once weekly GLP-1RA semaglutide reduced alcohol craving^22^. Similarly, GLP1-RA exenatide once weekly for 26 weeks reduced brain response, as measured by functional magnetic resonance imaging (fMRI), to alcohol cues in the ventral tegmental area^19^.

Although GLP-1RAs are known to have action in the brain, it is possible one mechanism by which GLP-1RAs reduce consumption of ingested drugs such as alcohol is, at least in part, through peripheral mechanisms.

While medications that reduce alcohol intake such as naltrexone and acamprosate have central nervous system action, disulfiram reduces alcohol intake through peripheral mechanisms.

Semaglutide, liraglutide and other GLP-1RAs have been shown to slow gastric emptying ^25–43^. Slowed gastric emptying can lead to a slower rise in blood alcohol, as alcohol is not well absorbed in the stomach. This can alter alcohol’s addictive potential as the pharmacokinetics of a substance is known to influence intake ^44–50^. Here, we sought to test these hypotheses by recruiting a cohort with obesity^19^ either taking GLP-1 RAs or not and test breath alcohol content (BrAc) and subjective feelings after an alcohol challenge dose. We observed that despite both groups consuming a dose of alcohol calculated to raise blood alcohol to 0.1 g/dl (approximately 0.08 g/dl BrAC)^51,52^, participants in the GLP-1RA group showed a slowed rise in BrAC and altered the subjective effects of alcohol.

## Methods

### Participants

Data were collected between July 2023 and May 2024. Participants (N = 20) were recruited from Roanoke, VA, and surrounding areas using flyers, the internet, and word-of-mouth referrals. To be eligible, participants had to occasionally drink alcohol, be 21 years of age or older, and have a BMI of 30 or greater. For the experimental group, participants had to be on a maintenance dose (at least 4 weeks, Supplemental Table 1) of a GLP-1RA medication for weight loss. Exclusion criteria were a history of bariatric surgery, alcohol use disorder identification test (AUDIT) greater than 16, hemoglobin A1c measurement greater than 9, a history of seizure disorder or traumatic brain injury, on specific medications that interfere with alcohol metabolism, or currently pregnant or lactating. All procedures were reviewed and approved by the Virginia Tech Institutional Review Board (#23-460).

### Measurement details

*Subjective feeling rating.* Participants were asked to rate their feeling of intoxication on a scale of 0-10, with 0 rated as not being drunk at all and 10 as the most drunk they have ever felt, by asking “How drunk do you feel right now?”

*Appetite score.* Participants were asked to indicate their current appetite using visual analog scales (VAS) as described by Flint and colleagues^78^. The VAS is a visual scale with each end representing the most positive and most negative ratings of hunger, satiety, fullness, and prospective food consumption. Appetite score was calculated as the average of each parameter.

*Alcohol urges questionnaire.* Craving was assessed using the Alcohol Urge Questionnaire (AUQ)79. Participants rate their feelings and thoughts about drinking in an 8-statement questionnaire at the time the measure is administered. Each question is a 7-item Likert scale ranging from “strongly disagree” to “strongly agree.” Craving score was calculated by averaging each item’s rating, with higher scores indicating greater craving.

*Nausea rating.* Participants rated their nausea on VAS anchored at No Nausea and Unbearable Nausea and prompted with “At this moment, how nauseous do you feel?”

*The Biphasic Alcohol Effects Scale (BAES).* BAES is a self-report, unipolar adjective rating scale that is designed to measure both stimulant and sedative effects of alcohol^80^. The scale consists of fourteen items that are rated on a scale of 0 (not at all) to 10 (extremely). Participants rated the extent to which drinking alcohol produced these feelings at present. Scores are calculated as the sum of the items.

*Additional questionnaires.* Participants reported on the good effects, the bad effects, taste and liking of alcohol during the alcohol administration session. Specifically, the VAS statements were: a) good effects: At this moment, are you experiencing good effects from the drink? (0 = No Good Effect, 50 = Neutral, and 100 = Strong Good Effects) b) bad effects: At this moment, are you experiencing bad effects from the drink? (0 = No Bad Effect, 50 = Neutral, and 100 = Strong Bad Effects) c) At this moment, how does the drink taste? (0 = Bad, 50 = Neutral, and 100 = Good), and d) At this moment, my liking for the drink served to me is? (0 = strong disliking, 50 = neither like nor dislike, and 100 = strong liking).

*Blood glucose.* First, the participant’s finger was cleansed with an alcohol pad, allowed to dry, then punctured with a disposable lancet. Blood was then collected and glucose assessed using a Contour Next EZ Blood Glucose Meter and Counter Test Strips distributed by Ascensia Diabetes Care US, Inc (New Jersey, USA).

*Breath Alcohol.* First, participants rinsed their mouths with water. Next, they were instructed to take a deep breath and blow the whole breath out into the breath alcohol analyzer (ALCO-SENSOR III, Intoximeters, Missouri, USA).

*Drink Stimulus.* The alcoholic beverage content was calculated using Formula 14 from Brick^81^ for each participant to reach 0.1 g/dl blood alcohol content (0.08 g/dl BrAC)^51,52^. A target of 0.1 g/dl was utilized to allow for a 20% variance between blood alcohol content (BAC) and BrAC^51,52^ (need citation). The multifactorial dose determination accounted for (1) time spent drinking, (2) time post-consumption, (3) total body water, (4) sex, (5) age, (6) weight, (7) height, and (8) estimated alcohol elimination rate. Each drink was mixed at a 1:3 ratio of Tito’s Vodka to Juice (cranberry or orange).

### Procedures

Prior to enrollment, all participants completed an online pre-screening survey, fingerstick test for HbA1c, urine pregnancy test, as well as height and weight measurements to ensure eligibility.

The study was approved by the VT IRB, and informed consent was obtained from all participants prior to their participation in the study. All methods were performed in accordance with relevant guidelines.

*Session 1.* After informed consent was obtained, participants completed a Timeline Follow Back (TLFB) to measure drinking in the past 30 days, Patient Health Questionnaire (PHQ-9), and a medical interview which asked about current and past medication history and other medical history to ensure safe participation in the study, as reviewed by a study physician.

*Session 2.* A day prior to the drinking session, participants were reminded to come in without eating anything. All participant sessions started between 8-10 AM. Once participants arrived, they were given a snack (Clif-Bar, Indianapolis, USA) to standardize caloric intake and ensure a similar gastric content prior to alcohol administration. Before proceeding to the Research Bar, the participants’ blood pressure, pulse, breath alcohol concentration and blood glucose level were measured. The participants then completed questionnaires related to drinking behaviors as described in the questionnaire section above.

Ninety minutes after consuming the snack, the participant was moved to the Research Bar. The drinking period of the session lasted one hour and was divided into three phases. In each phase the participant was served the prepared alcoholic beverage to be consumed within 10 minutes. Following this, participants were given a 10-minute break, completed subjective questionnaires about their current state and experience, including Visual Analogue Scales (VAS) as described above. During the breaks, research personnel also measured BrAC. This was repeated for three drinks over one hour.

Upon completion of the drinking period, participants were moved to a recovery room where they remained for 4 hours to allow the alcohol consumed to be metabolized to 0.02g/dL BrAC, the criterion for release. Researchers measured BrAC every 30 minutes, fingerstick blood glucose levels were measured immediately following the drinking session and at hour 3. At hour 3 participants completed the subjective questionnaires. After 4 hours in the recovery room and a BrAC <0.02g/dL, the participant was released with Study Physician approval.

## Results

### Characteristics of participants

We recruited 24 individuals with obesity into this study. At the time of study start (July 2023), the only available RCT evidence demonstrated a reduction in alcohol intake in people with obesity^19^. The 24 individuals with obesity were taking a maintenance dose of GLP-1RA medication for weight loss (N = 14, GLP-1RA group) for at least 30 days, or not taking any medication for weight loss (N = 10, control group). Of the 14 in the GLP-1RA group, 1 individual was lost to follow up, and 3 were not invited based on being excluded for having gastric bypass surgery (Supplementary Figure 1). The final sample (N = 20) consisted of majority white females, with a mean age of ~36 years and mean BMI of ~38. AUDIT scores were matched between controls with the GLP-1RA group (Range 1-6). Reported average drinking days were 1-2 in the past 30 days with an average of 1-2 drinks per use for both groups.

### The GLP-1RA group has reduced breath alcohol concentration (BrAC) and subjective feeling ratings during an alcohol administration session

To understand the effects of GLP-1RAs on alcohol absorption and its subjective effects, we compared BrAC and subjective feelings of intoxication between groups. Results of repeated measures linear mixed effect models indicated a significant interaction between group and time for BrAC (χ^2^(7)=16.36, p=0.022, f=0.36). We observed an initial blunted response at time points 10 (B=0.0163, p=0.013), 15 (B=0.0151, p=0.028) and 20 minutes (B=0.0196, p=0.001) in the GLP-1RA group as compared to the control group. From 35 minutes to 60 minutes, the GLP-1RA group’s BrAC was not significantly different than that of the control group. In addition, a main effect of sex (χ^2^(1) = 6.382, p=0.012, f=0.67) was observed with males having higher BrAC, overall. Additionally, cumulative BrAC calculated using area under the curve (AUC) was reduced in the GLP-1RA group as compared to the control group ((F(1,14)=12.41, p=0.003, f=0.94); inset, Fig 1A). The reduction indicates that the GLP-1RA medication delayed the initial rise in BrAC levels and resulted in more time with lower BrAC during the drinking session.

**Figure 1.**
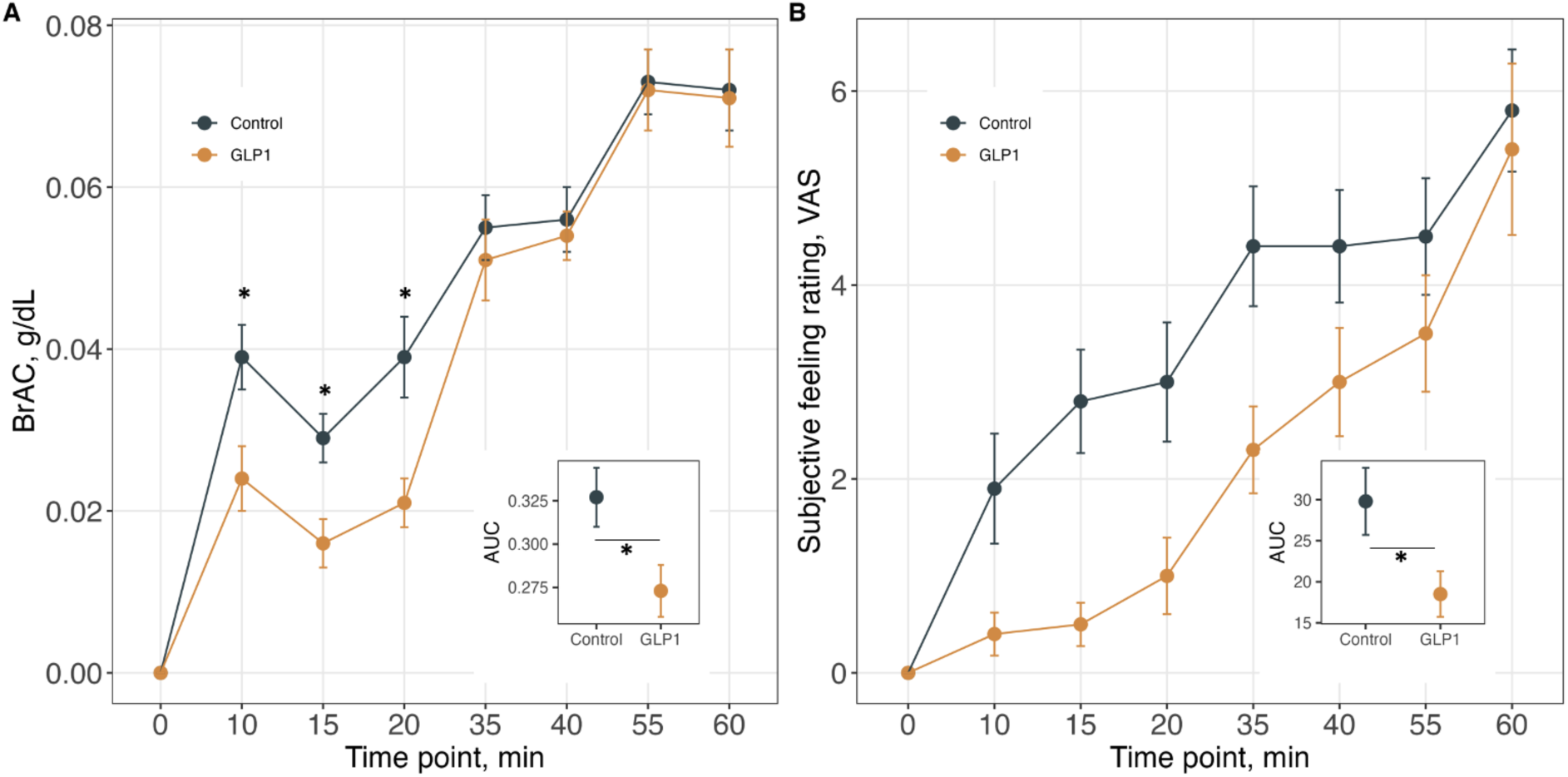
Objective and Subjective effects of GLP-1RAs on alcohol intoxication during an alcohol administration session. A) Breath alcohol concentration (BrAC) across drinking sessions. The x-axis displays timepoints (minutes), and the y-axis represents BrAC values (g/dL) during the drinking session. The inset shows the area under the curve reported during the 60 minutes. B) Subjective feeling ratings-VAS (“How drunk do you feel?”) across timepoints in the drinking session. The inset shows the area under the curve reported during the 60 minutes. * = p value < 0.05. Error bars represent standard errors. Group: Black – control, Gold – GLP-1RAs.

The GLP-1RA group reported lower subjective feelings of intoxication during the early timepoints of the session, as indicated by reduced ratings on the Visual Analog Scale (VAS) for “How drunk do you feel?”. A significant interaction between group and time (χ^2^(7)=18.85, p=0.008, f=0.39) was found in a linear mixed effects model, revealing that the GLP-1RA group consistently rated their intoxication lower across multiple timepoints compared to the control group (Figure 1B). Additionally, we calculated AUC for ratings across timepoints. The AUC was reduced in the GLP-1RA group as compared to the control group ((F(1,14)=4.21 p=0.05, f=0.55); inset, Fig 1B). This aligns with the objective BrAC measurements, demonstrating consistency between subjective experiences and physiological effects. Accordingly, BrAC was correlated with subjective feeling ratings in all participants regardless of group (χ^2^=114.4, p<0.001, f=0.90).

### Effect of GLP-1RAs on appetite and alcohol craving before and after an alcohol administration session

GLP-1RAs are known to reduce appetite and have been shown to reduce alcohol craving. Drinking alcohol increased appetite score in the control group, but not in the GLP-1RA group (Group x Time interaction (χ^2^(1) = 5.9106, p = 0.015, f = 0.57; Figure 2; Supplemental Table 1), indicating that the pattern of change in appetite before and after the drinking session differed between groups. Post hoc comparisons revealed a significant reduction in appetite scores in the control group from pre- to post-alcohol administration (B = −23.27, p = 0.0005), while the GLP-1RA group showed no significant change (B = −7.30, p = 0.41). Although the mean GLP-1RA group appetite score appeared higher, scores did not differ significantly between the control and GLP-1RA groups (p = 0.16).

**Figure 2.**
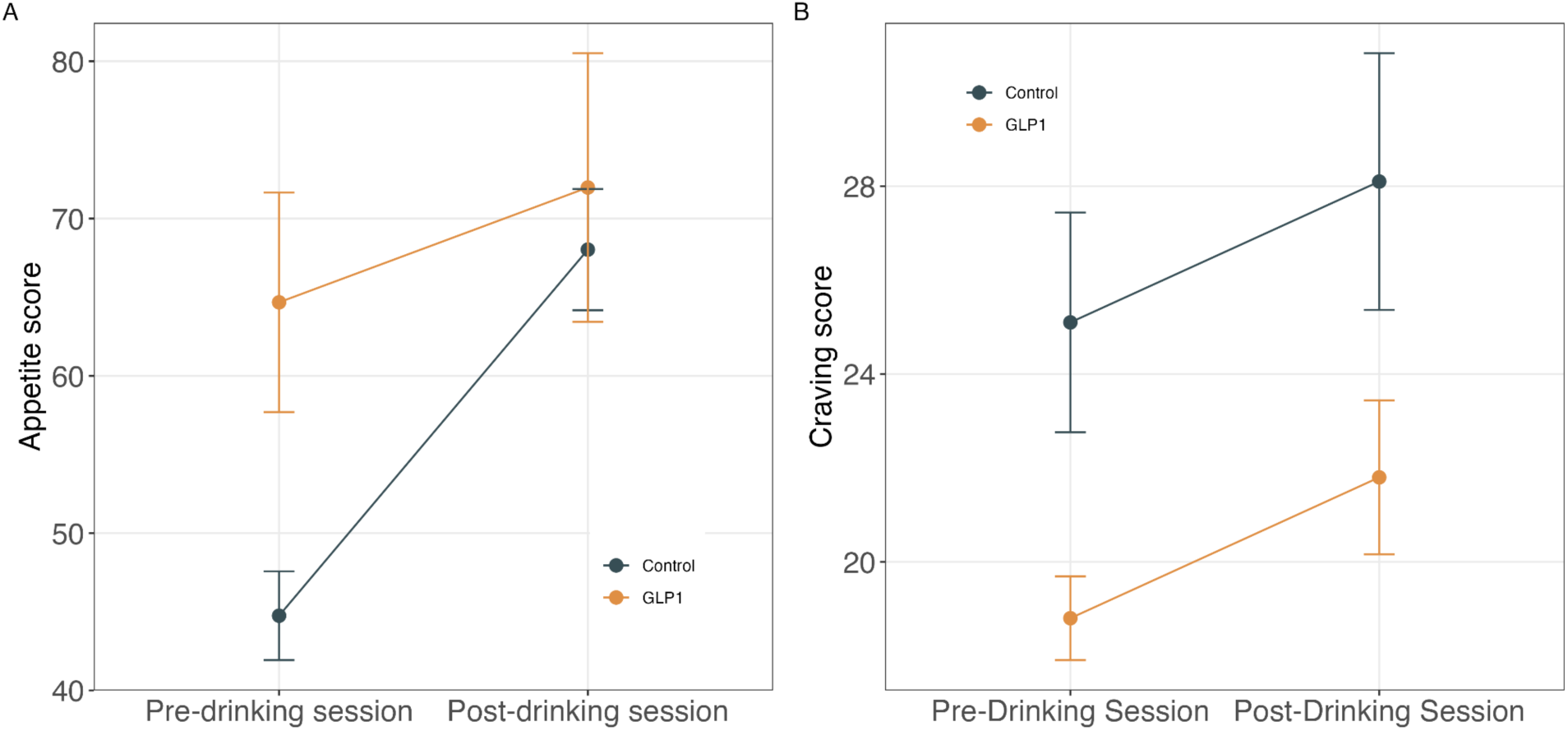
Appetite score and craving score before and after an alcohol administration session. A) Average appetite score between groups across the drinking session. The x-axis represents timepoint and the y-axis represents appetite scores. B) Average PACS score between groups across timepoints. The x-axis represents time point, and the y-axis displays the craving score. Error bars represent observed standard errors. Group: Black – control, Gold – GLP-1RAs. PACS: Penn Alcohol Craving Scale.

**Figure 3.**
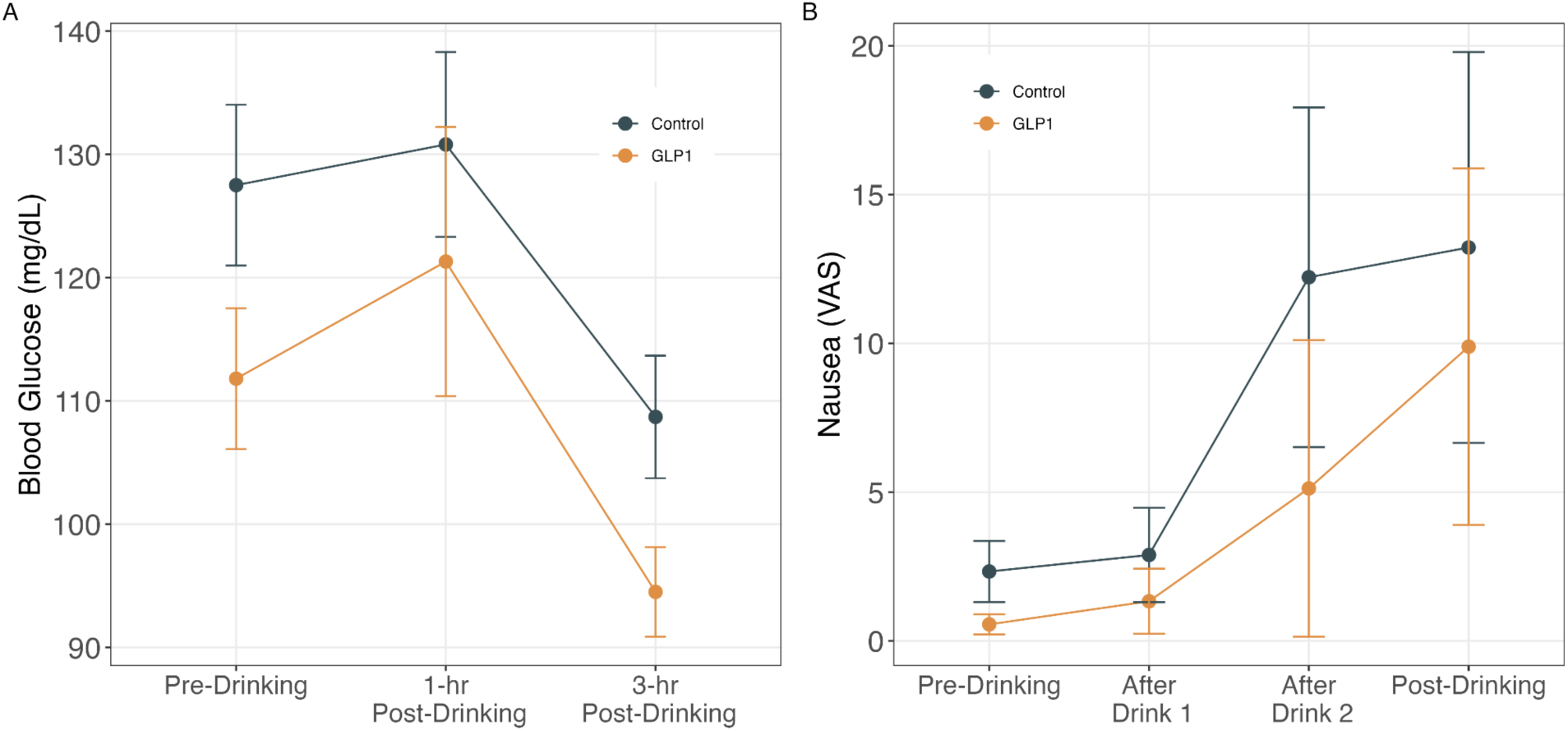
Blood glucose levels and nausea ratings before and after an alcohol administration session. A) Average blood glucose levels between groups across the drinking session. The x-axis displays pre- and post-drinking timepoints, and the y-axis represents blood glucose levels. B) Average nausea ratings between groups across the drinking session. The x-axis represents pre-, during, and post-drinking timepoints and the y-axis displays nausea scores. Error bars represent standard errors. Group: Black – control, Gold – GLP-1RA.

**Table 1.** Participant characteristics. Demographic and medication information of our sample. We observed no differences in these variables between the control and GLP-1RA groups.

|  | Overall | Control | GLP-1RA | p |
| --- | --- | --- | --- | --- |
| n | 20 | 10 | 10 |  |
| Age (mean (SD)) | 36.35 (9.85) | 34.90 (11.26) | 37.80 (8.56) | 0.525 |
| Education (mean (SD)) | 16.25 (2.94) | 16.00 (3.74) | 16.50 (2.01) | 0.714 |
| Employment (%) |  |  |  | 0.126 |
| Full Time | 14 (70.0) | 5 (50.0) | 9 (90.0) |  |
| Part Time | 2 (10.0) | 2 (20.0) | 0 (0.0) |  |
| Unemployed | 4 (20.0) | 3 (30.0) | 1 (10.0) |  |
| Ethnicity = Not Hispanic or Latino (%) | 19 (95.0) | 9 (90.0) | 10 (100.0) | 1.000 |
| Sex = M (%) | 4 (20.0) | 3 (30.0) | 1 (10.0) | 0.576 |
| Race = White (%) | 20 (100.0) | 10 (100.0) | 10 (100.0) |  |
| AUDIT_C (mean (SD)) | 3.55 (1.50) | 3.60 (1.71) | 3.50 (1.35) | 0.886 |
| BMI (mean (SD)) | 38.60 (5.33) | 37.98 (4.57) | 39.21 (6.18) | 0.619 |
| HbA1c (mean (SD)) | 5.22 (0.30) | 5.28 (0.32) | 5.16 (0.29) | 0.389 |
| Medication (%) |  |  |  |  |
| Liraglutide | 2 (20.0) | 0 (NaN) | 2 (20.0) |  |
| Semaglutide | 6 (60.0) | 0 (NaN) | 6 (60.0) |  |
| Tirzepatide | 2 (20.0) | 0 (NaN) | 2 (20.0) |  |
| Days on maintenance dose (mean (SD)) | 76.30 (55.07) | NaN (NA) | 76.30 (55.07) |  |

Next, we investigated the effects of GLP-1RAs on alcohol craving, measured before and after alcohol administration using PACS. Craving scores were significantly lower in the GLP-1RA group compared to the control group, as evidenced by a significant main effect of group (χ^2^(1) = 4.71, p = 0.03, f = 0.58; Supplemental Table 1). Additionally, craving scores increased after alcohol consumption across both groups, as shown by a significant main effect of time (χ^2^(1) = 6.15, p = 0.013, f = 0.57). However, no significant interaction of group by time was observed, indicating that GLP-1RAs did not differentially modulate the increase in craving across the drinking session.

### Effect of GLP1-RAs on blood glucose and nausea ratings before and after an alcohol administration session

Given GLP-1RAs are effective anti-glycemic medications, we examined whether alcohol consumption resulted in differential effects on blood sugar between groups. Blood glucose levels were significantly higher in the control group, with a significant main effect of group (χ^2^(1) = 2.84, p = 0.09, f = 0.45; Supplemental Table 1). Blood glucose for both groups changed over time, with a significant main effect of time point (χ^2^(1) = 15.09, p < 0.001, f = 0.63; Supplemental Table 1), but a significant interaction between time and group was not observed.

To rule out the possibility that the reduced subjective feelings of intoxication in the GLP-1RA group were due to nausea, we assessed nausea levels using a VAS (“How nauseous do you feel at this time?”) right after each drink during the alcohol administration session. For nausea, a significant main effect of time was observed (χ^2^(3) = 13.80, p = 0.003, f = 0.59; Supplemental Table 1), suggesting that nausea levels increased over the drinking session in both groups independently of group. Neither the Group effect nor an interaction between group and time reached statistical significance, indicating that the GLP-1RA group did not experience significantly different levels of nausea compared to the control group.

## Discussion

Here, we report that the acute physiological and subjective effects of alcohol differ in people with obesity taking a GLP-1RA from people with obesity not on a GLP-1RA medication. Specifically, after a challenge dose of alcohol, participants in the GLP-1RA group have a delayed rise in BrAC and a similar delayed rise in the subjective effects of alcohol. This difference in subjective effects was not explained by an increase in nausea in the GLP-1RA group. Participants in the GLP-1RA group reported lower craving of alcohol overall, although craving did rise over the course of the drinking session, as in the control group. This is consistent with previous studies reporting reduced alcohol craving after chronic GLP-1RA semaglutide administration^22^.

GLP1-RAs have been shown to have action in the central nervous system that mediates their weight loss and appetite suppressant effects ^53,54^. Accordingly, studies in rodents have demonstrated the ventral tegmental area and nucleus accumbens mediate the effects of GLP-1RAs on alcohol intake^11,12,55^. Fluorescently labeled semaglutide, one of the GLP1-RAs used in our study population, has been shown to bind in the nucleus accumbens^10^. In humans, 26 weeks of semaglutide reduced brain response to alcohol cues in the ventral tegmental area^19^. However, while central pharmacodynamics is an important factor influencing drug intake, peripheral pharmacokinetics can also have a profound effect on intake and even neural response to the same drug ^56,57^. This is most dramatically illustrated through differences in the intake patterns of the same drug with different routes of administration^45–50,58^, known as the “rate hypothesis” of addiction^49,59,60^.

One mechanism by which the rate of absorption of alcohol could be altered is through peripheral effects on gastric emptying in people taking GLP-1RAs. At least seventeen GLP-1RA clinical trials documented a delay in gastric emptying^25–41^. Furthermore, there have been multiple case reports of retained stomach contents or regurgitation following approved fasting protocols for patients on GLP-1RAs^61–71^. Together, these data indicate gastric emptying is slowed in people taking GLP1-RAs^29,42,72,73^. Alcohol is not readily absorbed in the stomach, but rather in the upper intestine^74^ and slowed gastric emptying would therefore slow the effects of alcohol.

Here, we find support for this hypothesis, in that our participants taking GLP1-RAs had a delayed rise in BrAC up to 20 minutes post alcohol consumption. This corresponded with a delayed increase in subjective feeling after alcohol consumption. We did not observe a difference in blood glucose changes in response to our alcohol challenge drink, which contained sugars in both the vodka and mixers. This could be taken as evidence that gastric emptying was similar for both groups, however, we obtained blood glucose levels 1 and 3 hours post drinking. BrAC levels in the GLP-RA group were not different from controls by 1 hour post drinking. Later studies should sample blood sugars more frequently to determine if the blood sugars and BrAC effects mirror each other.

## Limitations

While participants were matched between the GLP-1RA and control groups, our sample size was small. We have therefore included a table of all our statistical analyses with effect sizes as a reference for power analyses for future studies. While most of our participants were taking semaglutide, some were on liraglutide (n=2) and tirzepatide (n=2), and some received their medications through compounding pharmacies. We were not able to compare differences between drug type or source, though we have previously reported large reductions in subjective effects of alcohol in people taking tirzepatide^18^. This was not a randomized control trial, rather we recruited participants already taking these medications. Finally, while this study cannot definitively differentiate between central nervous system and peripheral effects of GLP-1RAs on alcohol intake, it does provide evidence that there is a role for peripheral mechanisms, which should be examined in future studies.

## Conclusions

With recent recommendations towards reductions in drinks as effective goals for individuals who use alcohol ^75–77^, these findings facilitate the consideration of GLP-1RAs for reducing alcohol use. While this study is small, we provide essential preliminary data (e.g., effect sizes) for the design and development of larger randomized control trials testing the effectiveness of GLP-1RAs on reductions in alcohol use.

## Acknowledgments

Funding was provided by Fralin Biomedical Research Institute at Virginia Tech Carillion. We would like to thank the participants for their time.

## Author Contributions

W.K.B, A.G.D., A.T., F.Q.: Conceptualization and design of study. F.Q., A.T., Z.E.: Data Analysis. M.F., F.Q., Z.E.,: Data Collection. F.Q., C.B., M.F., A.G.D.: Wrote the main manuscript text. A.T., A.G.D., K.G., A.K., W.K.B,: Supervision, Reviewing and editing the manuscript. F.Q., M.F., Z.E., K.G., A.K.: Recruitment, Administrative and IRB submission.

## Data Availability statement

The raw data is available from the corresponding author on reasonable request.

---

**Supplemental Figure 1.**
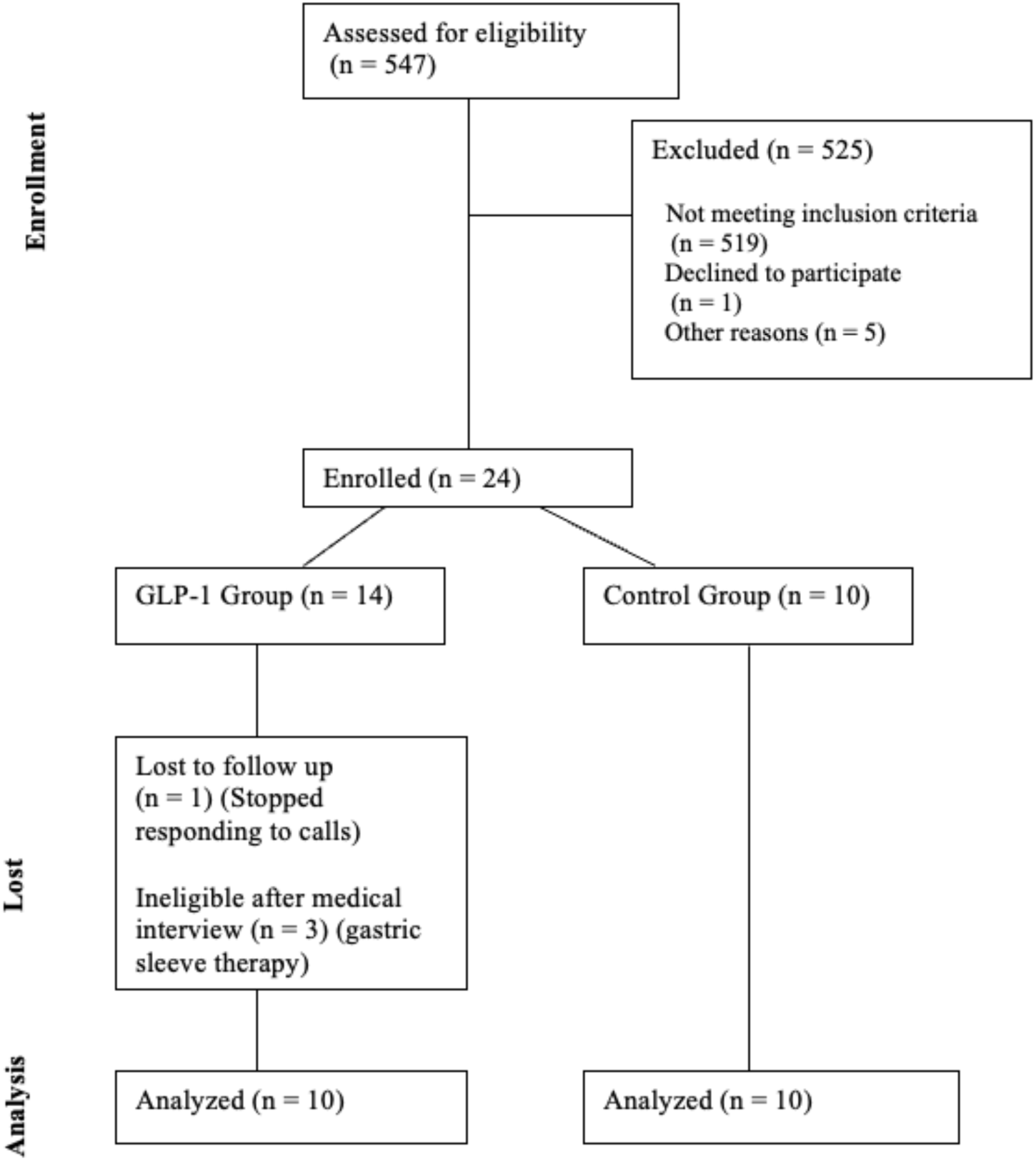
Consort Diagram.

**Supplemental Table 1.** Details about GLP-1RA Medications in our study. Medication, dose, and source, either compounding pharmacy or from the pharmaceutical company, are listed here for each participants in the GLP-1RA group.

| Participant | GLP-1RA | Days<br>(Maintenance<br>dose) | Dose | Source |
| --- | --- | --- | --- | --- |
| 1 | Tirzepatide | 140 | 15 mg | Pharmaceutical |
| 2 | Liraglutide | 190 | 1.8 mg | Pharmaceutical |
| 3 | Semaglutide | 35 | 2.4 mg | Pharmaceutical |
| 4 | Tirzepatide | 37 | 5 mg | Pharmaceutical |
| 5 | Liraglutide | 32 | 0.6 mg | Pharmaceutical |
| 6 | Semaglutide | 58 | 2 mg | Pharmaceutical |
| 7 | Semaglutide | 39 | 1 mg | Compounded |
| 8 | Semaglutide | 112 | 2 mg | Pharmaceutical |
| 9 | Semaglutide | 32 | 0.5 mg | Pharmaceutical |
| 10 | Semaglutide | 88 | 2.5 mg | Compounded |

