## Supplemental Table 2 for "Physiological and perceptual effects of GLP-1 receptor agonists during alcohol consumption in people with obesity: a pilot study"

| | BrAC | | Subjective | | Appetite | | Craving | | Sugar | | Nausea | |
|---|---|---|---|---|---|---|---|---|---|---|---|---|
| | Chi Sq. | Cohen's f | Chi Sq. | Cohen's f | Chi Sq. | Cohen's f | Chi Sq. | Cohen's f | Chi Sq. | Cohen's f | Chi Sq. | Cohen's f |
| Group | $\chi^2(1)$ = 0.112 | 0.031 | $\chi^2(1)$ = 0.037 | 0.034 | $\chi^2(1)$ = 4.835* | 0.516 | $\chi^2(1)$ = 4.710* | 0.580 | $\chi^2(1)$ = 2.846 | 0.451 | $\chi^2(1)$ = 0.711 | 0.275 |
| Time | $\chi^2(7)$ = 366.620*** | 1.706 | $\chi^2(7)$ = 184.498*** | 1.210 | $\chi^2(1)$ = 25.093*** | 1.181 | $\chi^2(1)$ = 6.151* | 0.569 | $\chi^2(2)$ = 15.094*** | 0.630 | $\chi^2(3)$ = 14.786** | 0.593 |
| Group:Time | $\chi^2(7)$ = 16.364* | 0.360 | $\chi^2(7)$ = 18.857** | 0.387 | $\chi^2(1)$ = 5.911* | 0.573 | | | | | | |
| Age | $\chi^2(1)$ = 1.706 | 0.349 | $\chi^2(1)$ = 0.000 | 0.004 | $\chi^2(1)$ = 0.017 | 0.035 | $\chi^2(1)$ = 0.734 | 0.229 | $\chi^2(1)$ = 0.135 | 0.098 | $\chi^2(1)$ = 0.001 | 0.403 |
| AUDIT_C | $\chi^2(1)$ = 0.613 | 0.209 | $\chi^2(1)$ = 1.518 | 0.329 | $\chi^2(1)$ = 0.53 | 0.195 | $\chi^2(1)$ = 2.327 | 0.408 | $\chi^2(1)$ = 0.988 | 0.266 | $\chi^2(1)$ = 2.247 | 0.753 |
| BMI | $\chi^2(1)$ = 1.994 | 0.377 | $\chi^2(1)$ = 0.032 | 0.048 | $\chi^2(1)$ = 1.843 | 0.363 | $\chi^2(1)$ = 0.248 | 0.133 | $\chi^2(1)$ = 0.006 | 0.020 | $\chi^2(1)$ = 0.910 | 0.035 |
| Sex | $\chi^2(1)$ = 6.382* | 0.675 | $\chi^2(1)$ = 1.057 | 0.275 | $\chi^2(1)$ = 0.543 | 0.197 | $\chi^2(1)$ = 2.026 | 0.380 | $\chi^2(1)$ = 0.575 | 0.203 | $\chi^2(1)$ = 0.052 | 0.196 |

| | BAES (Stimulative) | | BAES (Sedative) | | Good effect (VAS) | | Bad effect (VAS) | | Taste (VAS) | | Liking (VAS) | |
|---|---|---|---|---|---|---|---|---|---|---|---|---|
| | Chi Sq. | Cohen's f | Chi Sq. | Cohen's f | Chi Sq. | Cohen's f | Chi Sq. | Cohen's f | Chi Sq. | Cohen's f | Chi Sq. | Cohen's f |
| Group | $\chi^2(1)$ = 0.001 | 0.006 | $\chi^2(1)$ = 1.259 | 0.236778667 | $\chi^2(1)$ = 0.19 | 0.0981 | $\chi^2(1)$ = 0.23 | 0.105 | $\chi^2(1)$ = 0.221 | 0.111 | $\chi^2(1)$ = 1.743 | 0.311 |
| Time | $\chi^2(1)$ = 14.218*** | 0.889 | $\chi^2(1)$ = 0.8 | 0.210837335 | $\chi^2(2)$ = 7.633* | 0.4738 | $\chi^2(2)$ = 5.811* | 0.413 | $\chi^2(2)$ = 2.917 | 0.297 | $\chi^2(2)$ = 0.782 | 0.152 |
| Group:Time | $\chi^2(1)$ = 0.332 | 0.136 | $\chi^2(1)$ = 0.358 | 0.141025887 | $\chi^2(2)$ = 1.677 | 0.2216 | $\chi^2(2)$ = 2.448 | 0.268 | $\chi^2(2)$ = 0.687 | 0.144 | $\chi^2(2)$ = 0.996 | 0.171 |
| Age | $\chi^2(1)$ = 0.141 | 0.100 | $\chi^2(1)$ = 0.072 | 0.071710872 | $\chi^2(1)$ = 0.251 | 0.1346 | $\chi^2(1)$ = 0.007 | 0.022 | $\chi^2(1)$ = 0.029 | 0.045 | $\chi^2(1)$ = 0.192 | 0.117 |
| AUDIT_C | $\chi^2(1)$ = 0.174 | 0.111 | $\chi^2(1)$ = 2.21 | 0.397286267 | $\chi^2(1)$ = 0.001 | 0.0093 | $\chi^2(1)$ = 2.102 | 0.388 | $\chi^2(1)$ = 2.391 | 0.413 | $\chi^2(1)$ = 3.673* | 0.513 |
| BMI | $\chi^2(1)$ = 1.218 | 0.295 | $\chi^2(1)$ = 1.07 | 0.276400716 | $\chi^2(1)$ = 1.991 | 0.3792 | $\chi^2(1)$ = 0.202 | 0.121 | $\chi^2(1)$ = 0.365 | 0.162 | $\chi^2(1)$ = 0.01 | 0.027 |
| Sex | $\chi^2(1)$ = 7.187** | 0.716 | $\chi^2(1)$ = 0.01 | 0.027240238 | $\chi^2(1)$ = 0.015 | 0.0326 | $\chi^2(1)$ = 0.457 | 0.178 | $\chi^2(1)$ = 0.701 | 0.222 | $\chi^2(1)$ = 0.148 | 0.102 |

Supplemental Table 2. ANOVA and effect sizes for variables of interest. Here we report the detailed statistics and effect sizes of all our outcome variables. Bolded values are $p<0.05$. Effect sizes are reported to inform power analyses for future studies.
